# Nomograms for Predicting the Risk of Acute Heart Failure in ICU Patients with Atrial Fibrillation: A Retrospective Cohort Study

**DOI:** 10.1101/2023.12.05.23299546

**Authors:** Ziyang Wu, Yong Qiao, Gaoliang Yan, Yuhan Qin, Huihong Tang, Shiqi Liu, Dong Wang, Chengchun Tang

**Affiliations:** Department of Cardiology, Zhongda hospital, School of Medicine, Southeast University, Dingjiaqiao 87, Gulou district, Nanjing 210009, PR China

**Keywords:** nomogram, Medical Information Mart for Intensive Care, acute heart failure, atrial fibrillation

## Abstract

**Aim:** This study developed two nomograms to predict the incidence of acute heart failure (AHF) in patients of atrial fibrillation (AF) and evaluated the predictive value of the nomograms.

**Method:** 18179 patients of AF from the Medical Information Mart for Intensive Care IV (MIMIC-IV) database were analysed. The patients were randomly divided into two groups in a seven-to-three ratio to form a training cohort (n = 12725) and a validation cohort (n = 5454). Least absolute shrinkage and selection operator (LASSO) regression analyses were used to identify associated risk variables. Two nomograms were established to predict the rate of AHF in patients of AF for use by patients and clinicians, respectively. The new models were assessed in terms of the concordance index (C-index), the area under the curve (AUC) of receiver operating characteristic (ROC) analysis, calibration curve, and decision curve analysis (DCA).

**Results:** Least absolute shrinkage and selection operator (LASSO) regression analysis identified seven potential predictors of acute heart failure in nomogram 1 and three potential predictors in nomogram 2. Multivariate logistic regression analysis was used to evaluate the effects of these predictors and create final models. The concordance index values were 0.768 and 0.696, respectively. The areas under the curves for the training and validation sets in nomogram 1 were 0.768 and 0.763, respectively. The areas under the curves for the training and validation sets in nomogram 2 were 0.696 and 0.692, respectively.

**Conclusion:** The age, respiratory rate, fluid management, mean corpuscular hemoglobin concentration (MCHC), bicarbonate, blood urea nitrogen (BUN) and chloride were identified as predictors of the development of acute heart failure in patients with atrial fibrillation in nomogram 1. The charlson score, lods score and sapsII score were identified as predictors of the development of acute heart failure in nomogram 2. Our nomograms are reliable convenient approaches for predicting acute heart failure in patients with atrial fibrillation.

## Introduction

Atrial fibrillation (AF) is an atrial ectopic tachyarrhythmia [1]. The loss of conduction and refractory period synchronisation in various parts of the atrium leads to uncoordinated AF in muscle fibres [2]. Possible mechanisms for AF include ectopic pacing points, circular motion, and multiple micro re-entrances in the atrium [3, 4]. The Global Burden of Disease study has estimated that there are 33.5 million patients with AF [5]. AF leads to 2-fold and 1.5-fold increases in the all-cause mortality rate in women and men, respectively, due to progressive heart failure, cardiac arrest, and embolism; thus, it is a large burden on society [4, 6, 7].

The relationship between heart failure and AF has received increasing attention in recent years [8]. AF and acute heart failure (AHF) are increasing in prevalence, exacerbate each other, and share clinical features [9, 10]. Animal and human studies have confirmed the connection between AHF and AF [10–12]. The mechanisms include activation of the renin angiotensin aldosterone system (RAAS), mechanical traction on the left atrium, and atrial electrophysiological remodelling [10, 12, 13-15]. AF also plays a role in the occurrence and progression of AHF [16]. Thus far, research has mainly focused on the incidence of AF in patients with AHF; few studies have examined the incidence of AHF in patients with AF.

In oncology and pharmacy, nomograms are widely used to determine prognosis and evaluate the incidence of an event [17]. This study used data from the MIMIC-IV database to establish and validate two nomograms that can predict the incidence of AHF in patients with AF; these nomograms can be used by patients and clinicians, respectively.

## Methods

### Aim

This study aimed to determine the incidence of AHF in patients with AF.

### Database

MIMIC-IV is a publicly available database containing data from 76,540 intensive care unit (ICU) stays from 2008 to 2019 in the Beth Israel Deaconess Medical Center [18, 19]. This database contains all baseline data of patients and most laboratory results that can be used for various studies [20]. It aims to eliminate obstacles to scientific research [21]. The version we used is v2.2, and PostgreSQL v16 was used to search and extract data from it (http://www.postgresql.org/) [22].

### Study populations

This is a retrospective analysis using data from patients with AF stored in MIMIC-IV database. The inclusion criteria were as follows:

1. Patients with previous history or new onset of AF.
2. Patients aged older than 18 years.
3. Patients admitted to the ICU for the first time.

### Data collection and measurement

The characteristics used for our work included demographic data (e.g., age, gender, race and marital status), vital signs (e.g., heart rate, blood pressure and fluid management), laboratory results (e.g., routine blood tests, chemistry and coagulation) and common score systems in the ICU (e.g., LODS score, CHARLSON score and OASIS score). All data were collected six hours before admission to the ICU to 24 hours after admission to the hospital. Unmarried, married, divorced and widowed status was marked as 0, 1, 2 and 3, respectively. White people were marked as 0, and people of other races were marked as 1. Women were marked as 0, and men were marked as 1.

### Statistical analysis

Statistical analysis was achieved by using R software (Version 4.1.3; R Foundation for Statistical Computing, Vienna, Austria; https://www.r-project.org) [23]. The distribution of continuous variables was assessed using the Shapiro–Wilk test. Mean and standard deviation (SD) are reported for continuous variables having a normal distribution; median and interquartile range (IQR), for continuous variables having a skewed distribution; and numbers and percentages, for categorical variables. The Kruskal–Wallis H test was used to compare continuous data. The Fisher’s exact test was used to compare categorical variables. Seventy percent of the data were allocated to the training set for developing the model, and the remaining thirty percent were allocated to the validation set for validation. Records with missing values were deleted or replaced by using chained equations as appropriate. The LASSO method, which accounts for multicollinearity and avoids model overfit, was used to select the best candidate variable for establishing the nomograms [24]. To avoid making the clinical prediction models too complex, the lambda value was set to ‘1se’ so that the error was within one standard error of the minimum. Sensitivity and specificity were used to evaluate the models’ performance. The calibration C index (bootstrap resampling performed 1000 times), the calibration curve and ROC curve were used to evaluate the discrimination and calibration of the models [25]. DCA was used to identify the clinical benefit of the models [26]. The area under the receiver operating characteristic curve values were drawn to assess the stability of the models [27]. All significance tests were two-sided, and a P value < 0.05 was considered statistically significant.

## Results

### Clinical characteristics

Figure 1 is a flowchart showing the cohort selection process. This study included 18179 patients. Of these, 70% (12725) were randomly assigned to the training set and the remaining 30% (5454) were assigned to the validation set.

**Figure 1.**
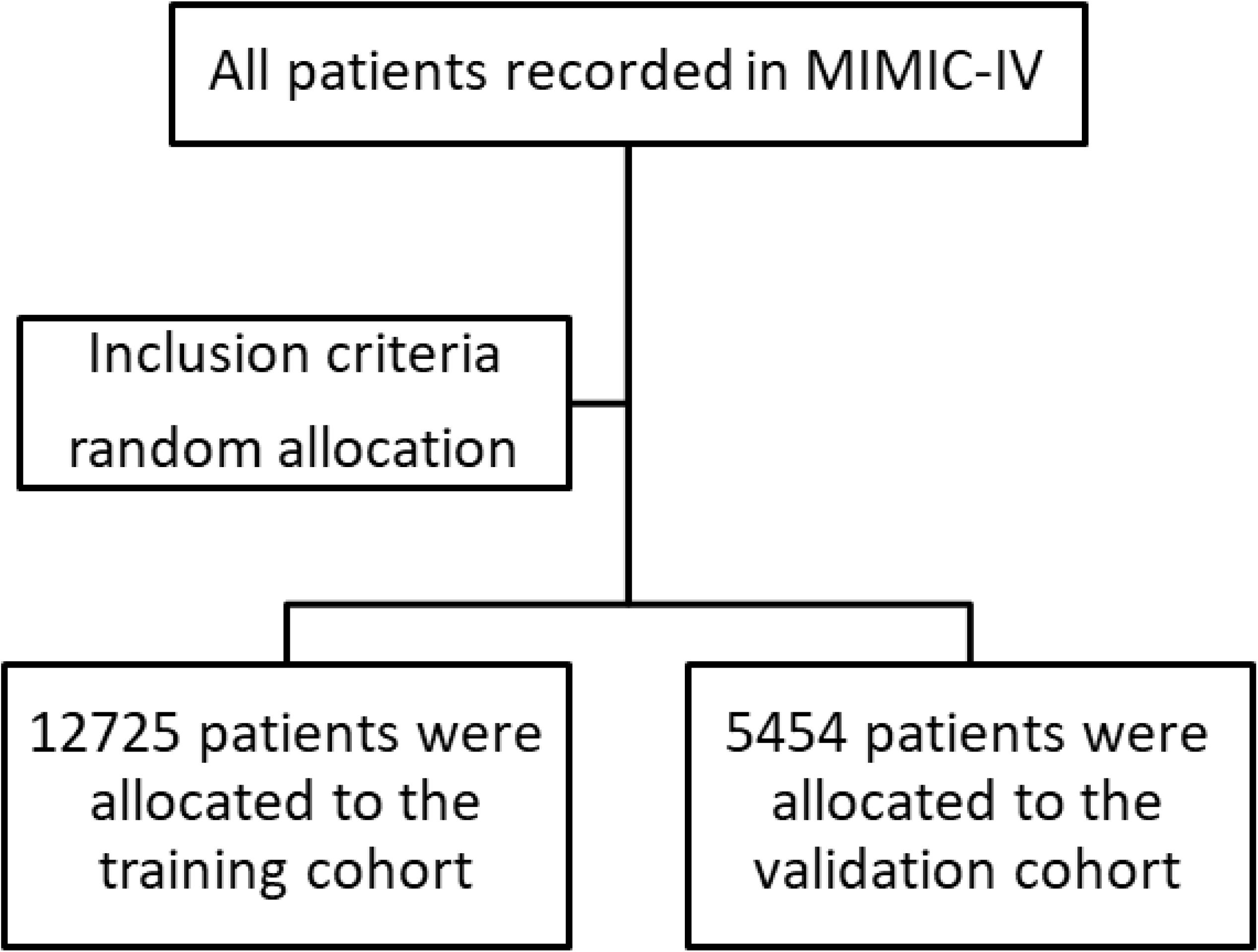
Flowchart showing the cohort selection process.

Table 1 summarizes the data of the enrolled patients. Of the 18179 patients with AF, 4582 were admitted to the ICU for an AHF. The age, weight, body mass index (BMI), heart rate, respiratory rate, fluid management, red blood cells (RBC), white blood cells (WBC), platelet, mean corpuscular volume (MCV), red blood cell distribution width (RDW), anion gap, bicarbonate, BUN, creatinine, Ca, blood glucose, Na, K, international normalized ratio (INR), prothrombin time (PT), partial thromboplastin time (PTT), apsⅢ score, charlson score, lods score and sapsⅡ score were higher in AHF patients with AF than in AHF patients without AF, the height and gcs score were the same, while the mean blood pressure, temperature, hemoglobin, hematocrit, mean corpsular hemoglobin (MCH), mean corpuscular hemoglobin concentration (MCHC), chloride, and RDW were lower. Widowed female patients of AF were more likely to develop AHF.

**Table 1.**
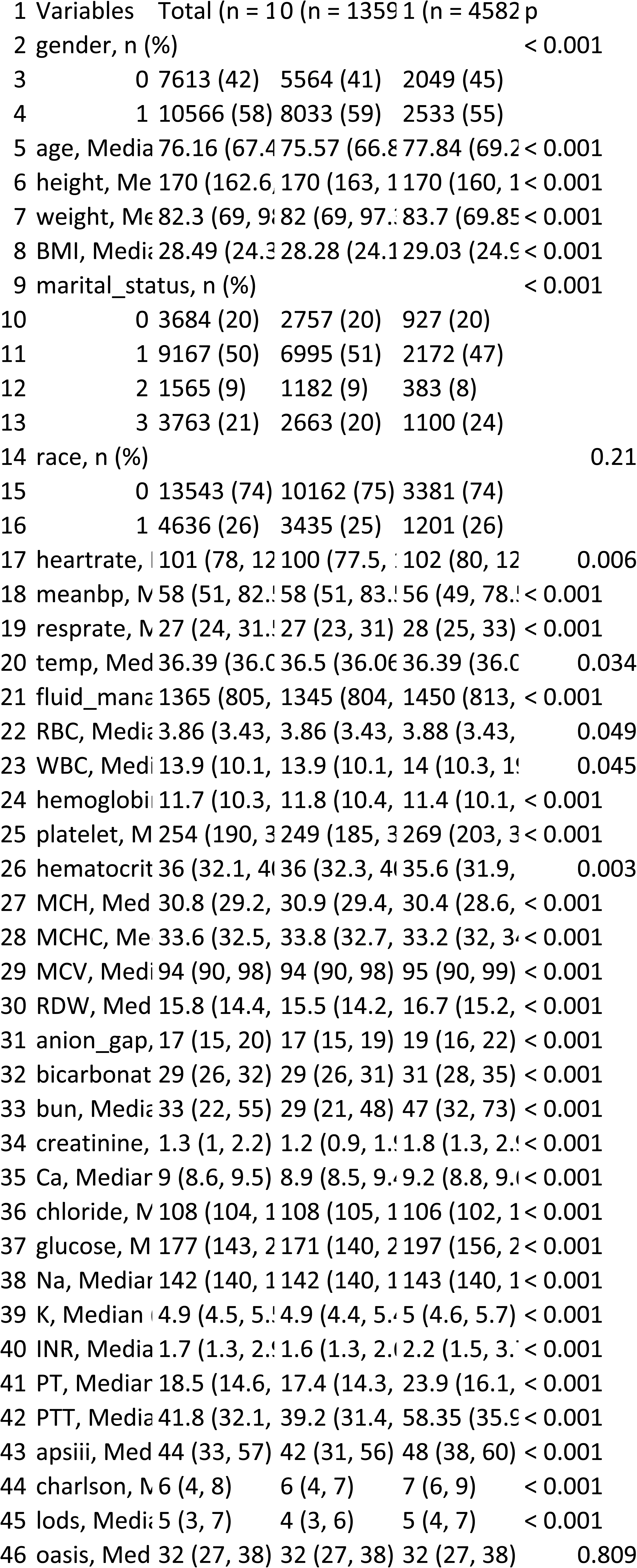
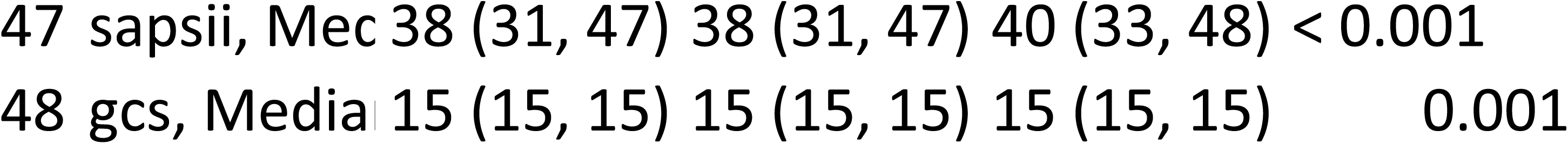
The data on patients enrolled in this study.

### Variable selection

Initially, 18189 patients and 46 variables were screened. After removing variables with missing values exceeding 5% of the total, 39 variables were retained in the study. Variables with missing values not exceeding 5% were replaced by using chained equations. The values of troponin-T and N-terminal pro-b-type natriuretic peptide were missing in more than 50% and 70% of patients, respectively. Although they are significant predictors of cardiac function, once these indicators are tested, it is possible to determine whether AMI has occurred, so these variables were excluded. Twentyfold cross-validated LASSO regression and manual screening identified ten variables: age, respiratory rate, fluid management, MCHC, bicarbonate, BUN and chloride for nomogram 1, and charlson score, lods score and sapsⅡ score for nomogram 2. Figures 2 and 3 illustrate the variable selection process. These ten variables were included in the multivariate logistic regression model for predicting the occurrence of AHF, and their odd ratios and 95% confidence intervals (CIs) were 1.32 (1.20–1.46), 1.16 (1.10–1.22), 1.34 (1.25–1.43), 0.72 (0.67-0.79), 1.78 (1.66-1.92), 1.89 (1.77-2.02), 0.69 (0.64-0.74), 2.61 (2.36-2.88), 2.34 (2.07-2.66) and 0.55 (0.49-0.63) respectively.

**Figure 2.**
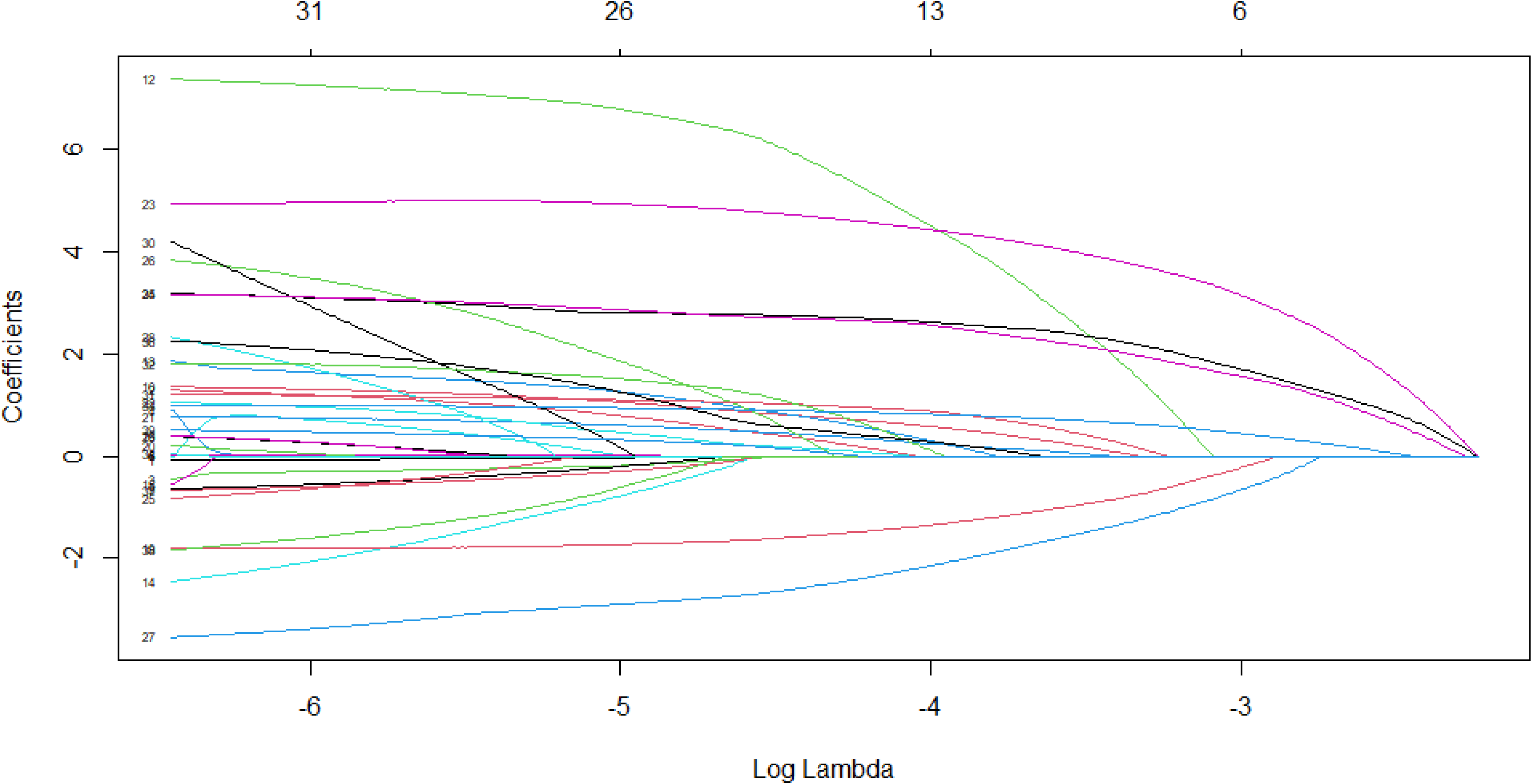
The variable selection process.

**Figure 3.**
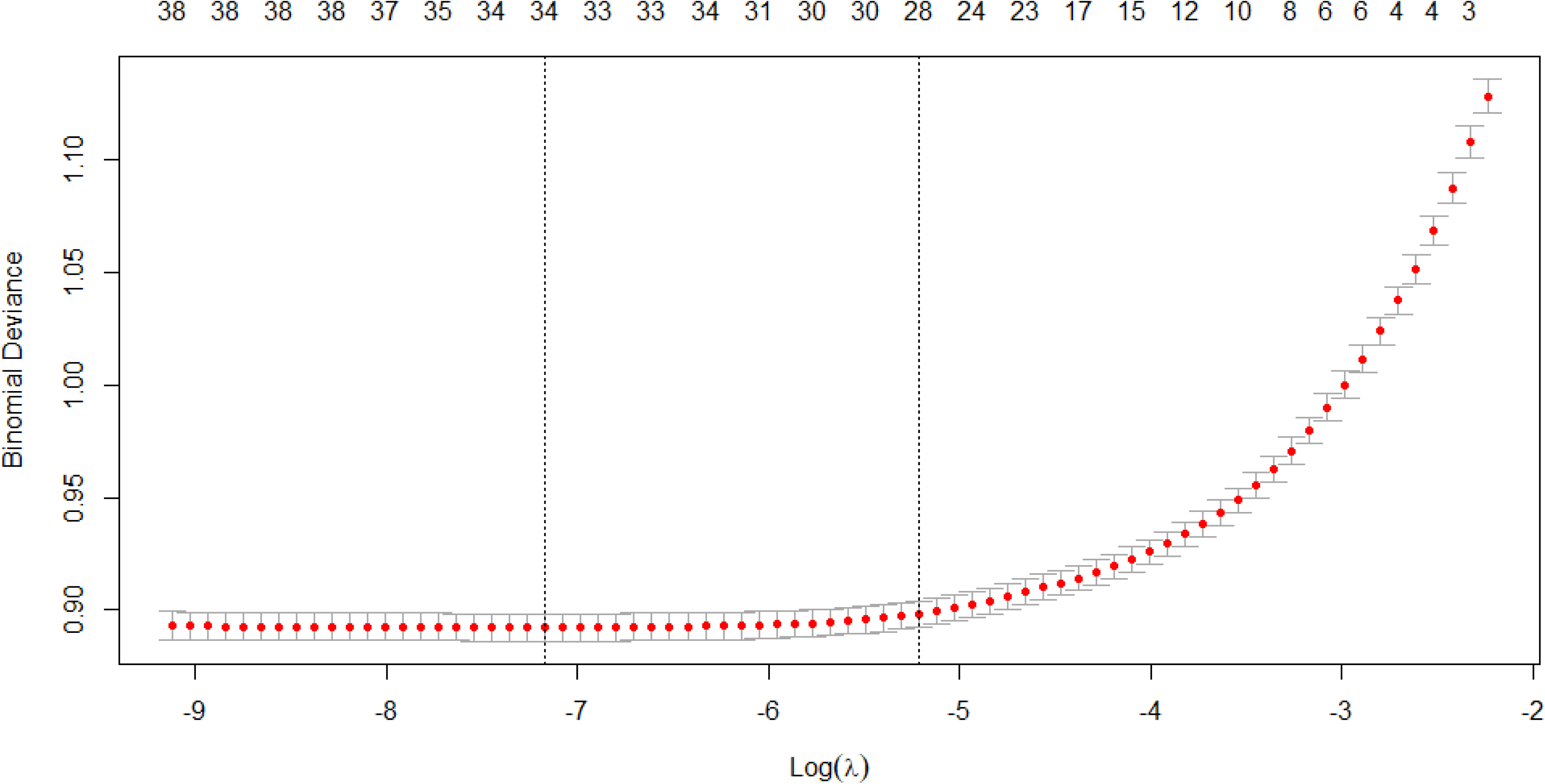
The variable selection process.

### Nomograms construction

A nomogram was established for predicting the occurrence of AHF among AF patients for use by patients (Figure 4) and the other nomogram was established for use by clinicians (Figure 5). The ten variables identified above were assigned different initial scores and a total score was calculated. The probability of developing AHF increased with the total score.

**Figure 4.**
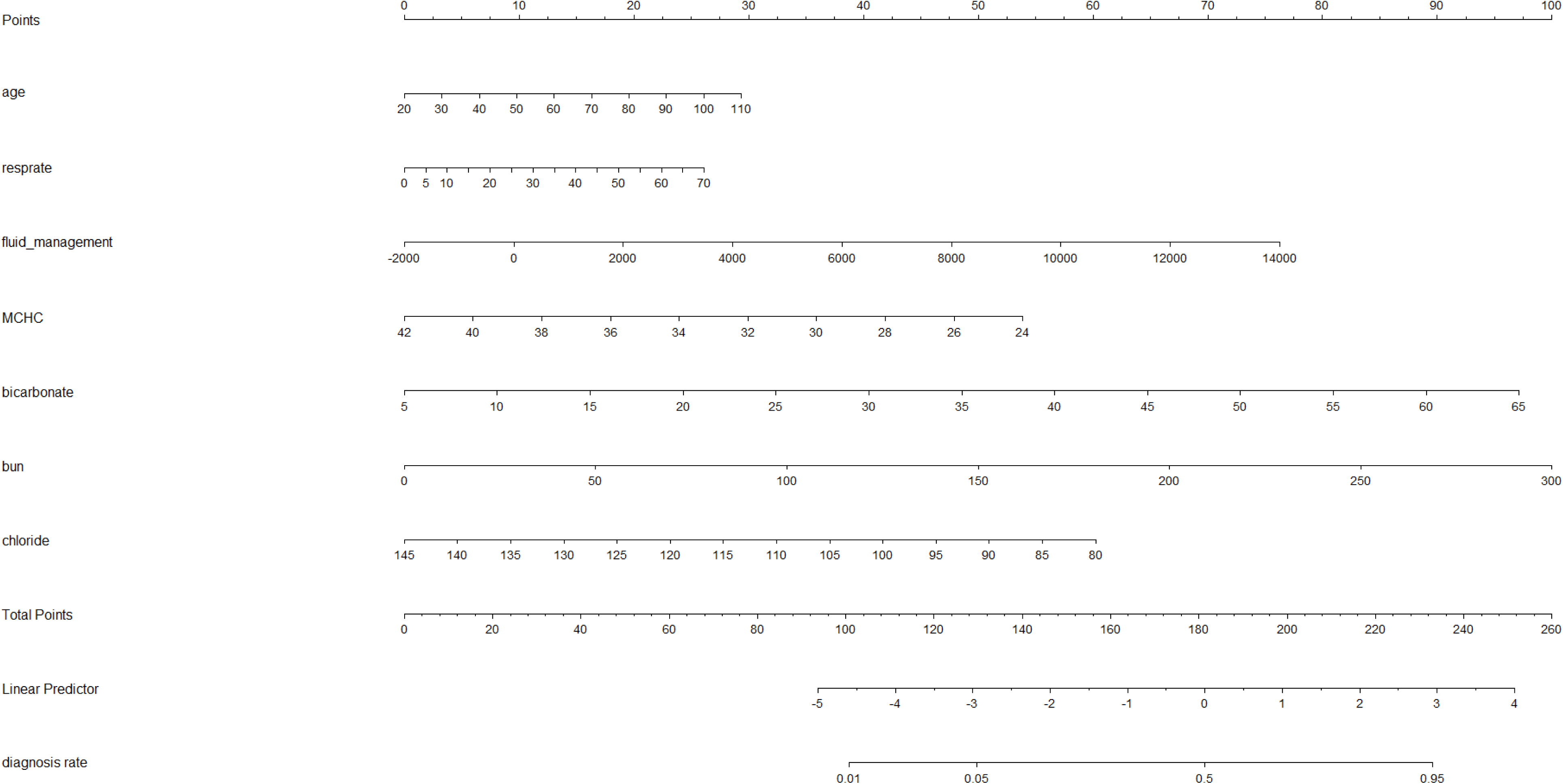
Nomogram for predicting the occurrence of AHF in AF patients.

**Figure 5.**
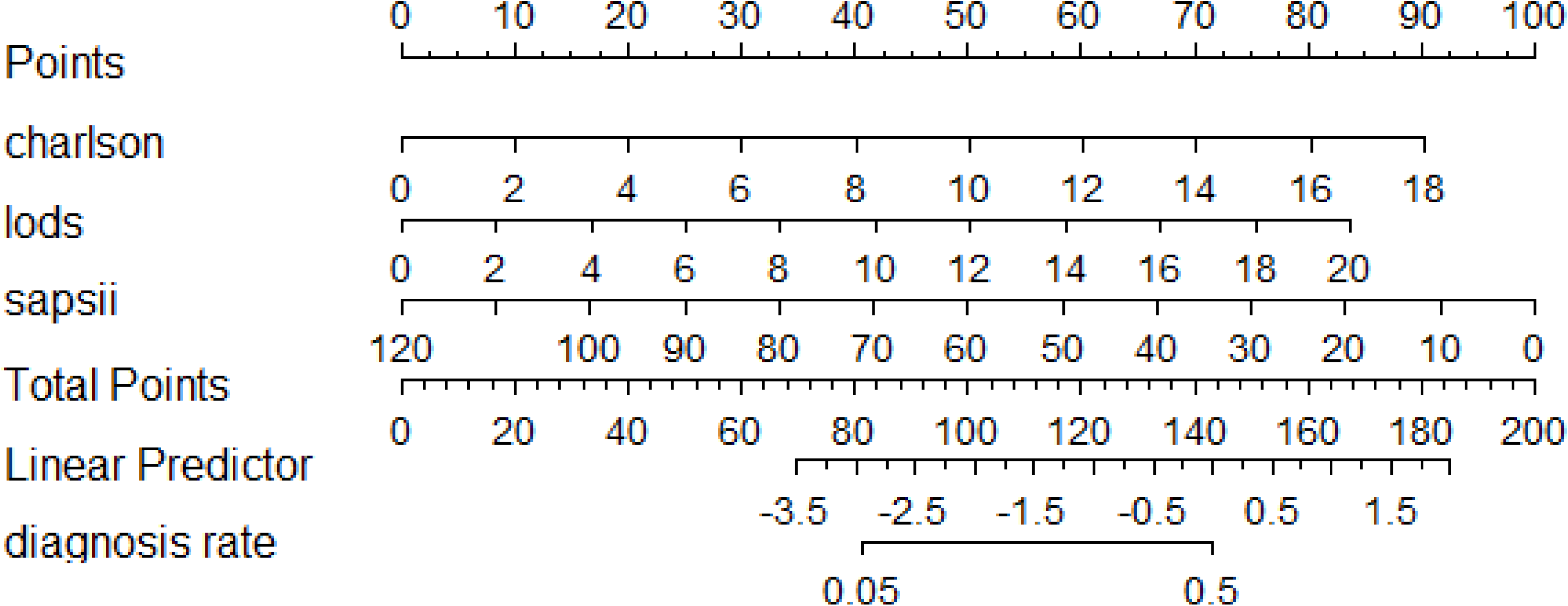
Nomogram for predicting the occurrence of AHF in AF patients.

### Assessment and validation

The calibration curves demonstrated high conformity between the predicted and observed probabilities (Figure 6 and 7). ROC curves for the training and test sets were used to evaluate the discriminability of the models. The area under the ROC curve was 0.768 in the training set and 0.763 in the validation set in nomogram 1 (Figures 8 and 9). The area under the ROC curve was 0.696 in the training set and 0.692 in the validation set in nomogram 2 (Figures 10 and 11). The results indicated that our prediction models have good stability and accuracy.

**Figure 6.**
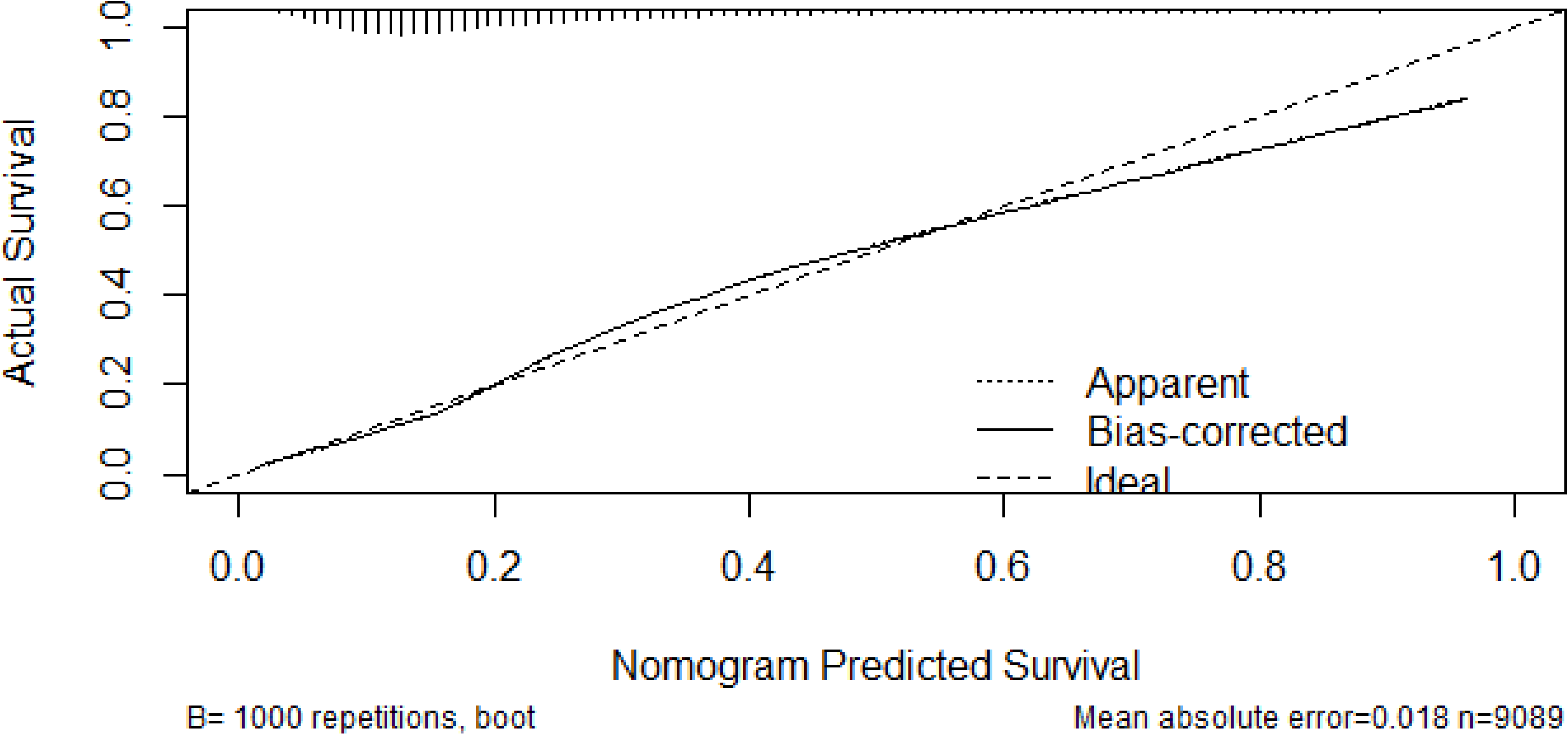
The calibration curve of nomogram 1.

**Figure 7.**
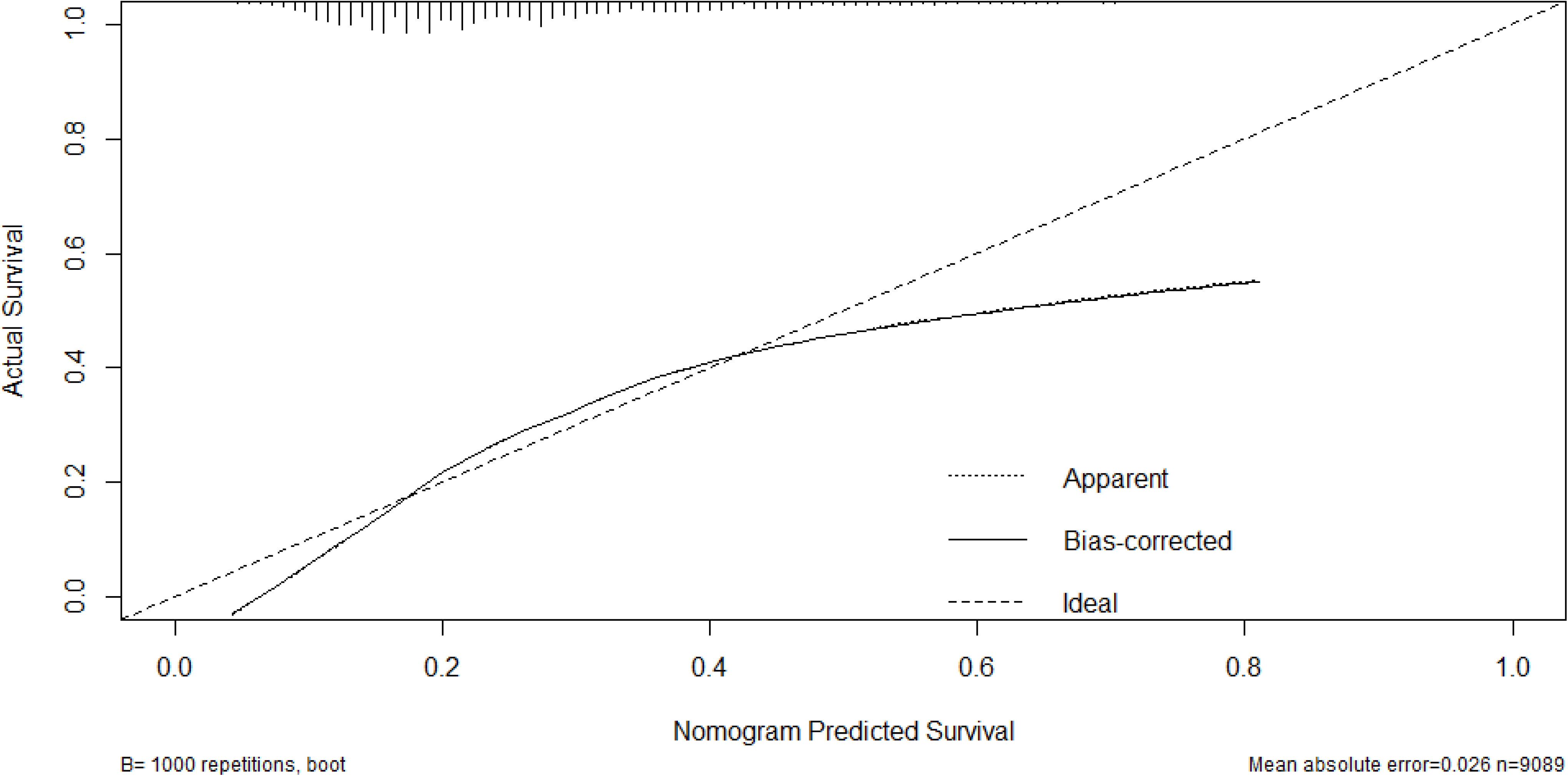
The calibration curve of nomogram 2.

**Figure 8.**
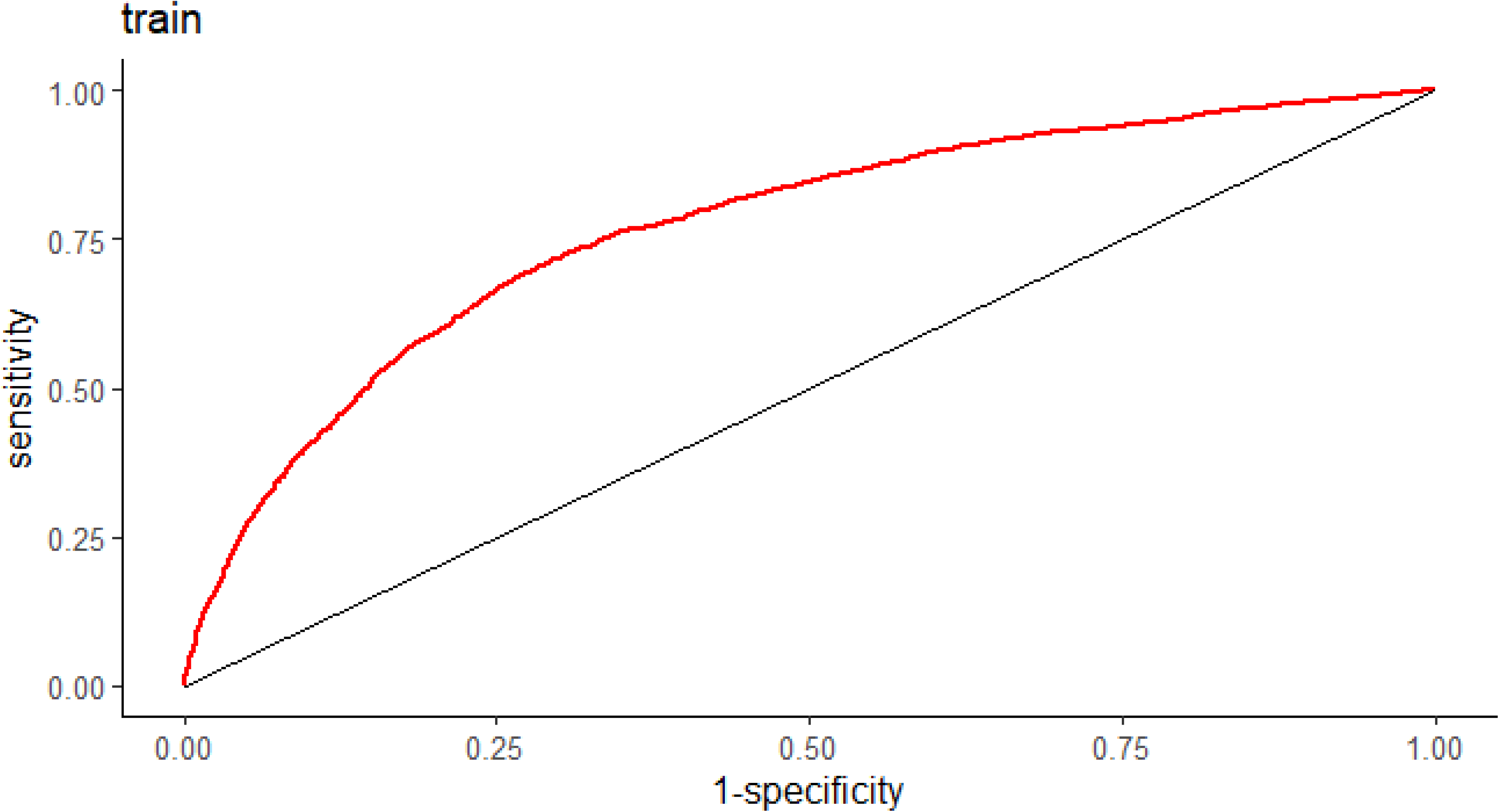
The ROC curve of the training set in nomogram 1.

**Figure 9.**
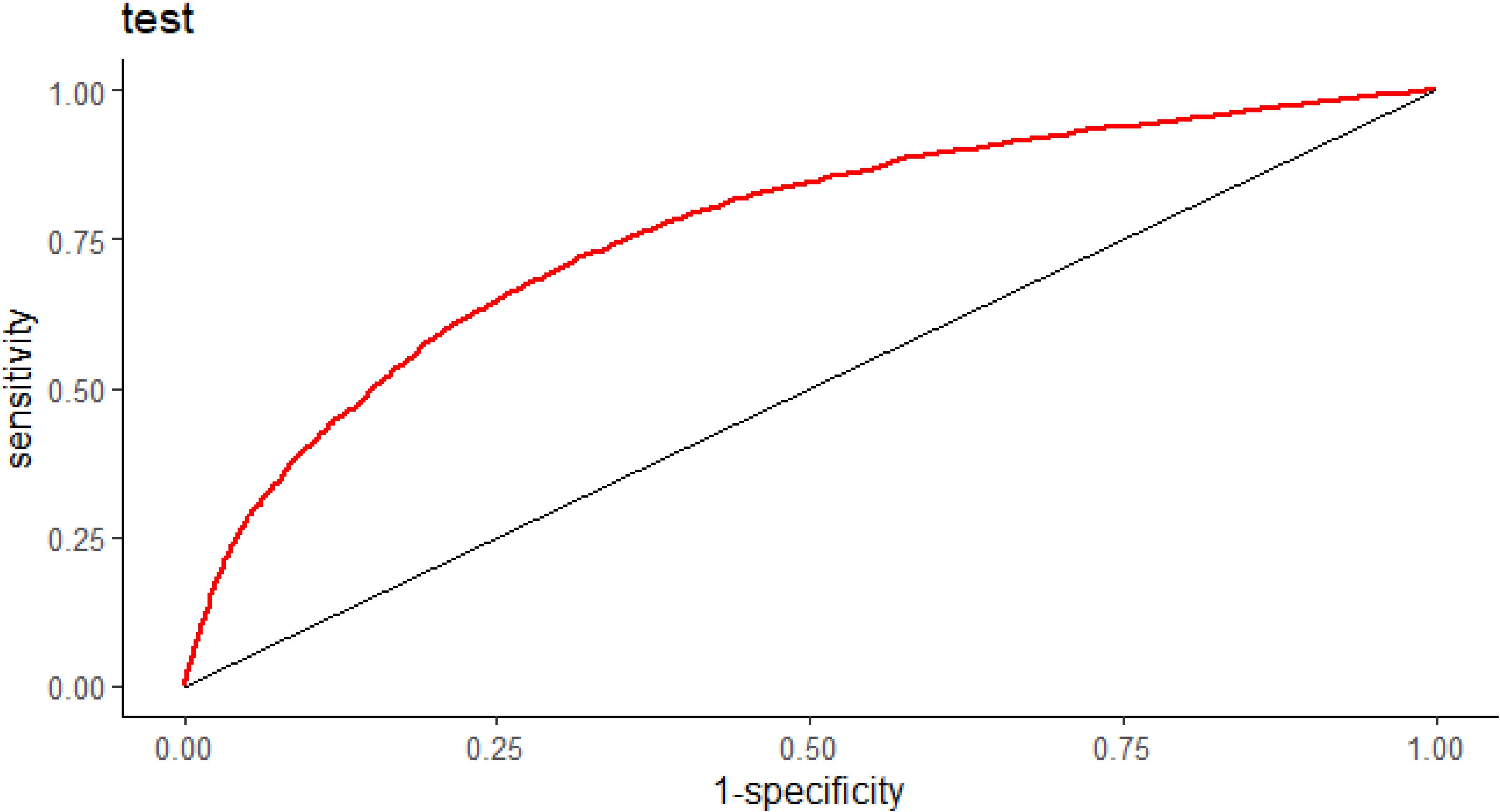
The ROC curve of the validation set in nomogram 1.

**Figure 10.**
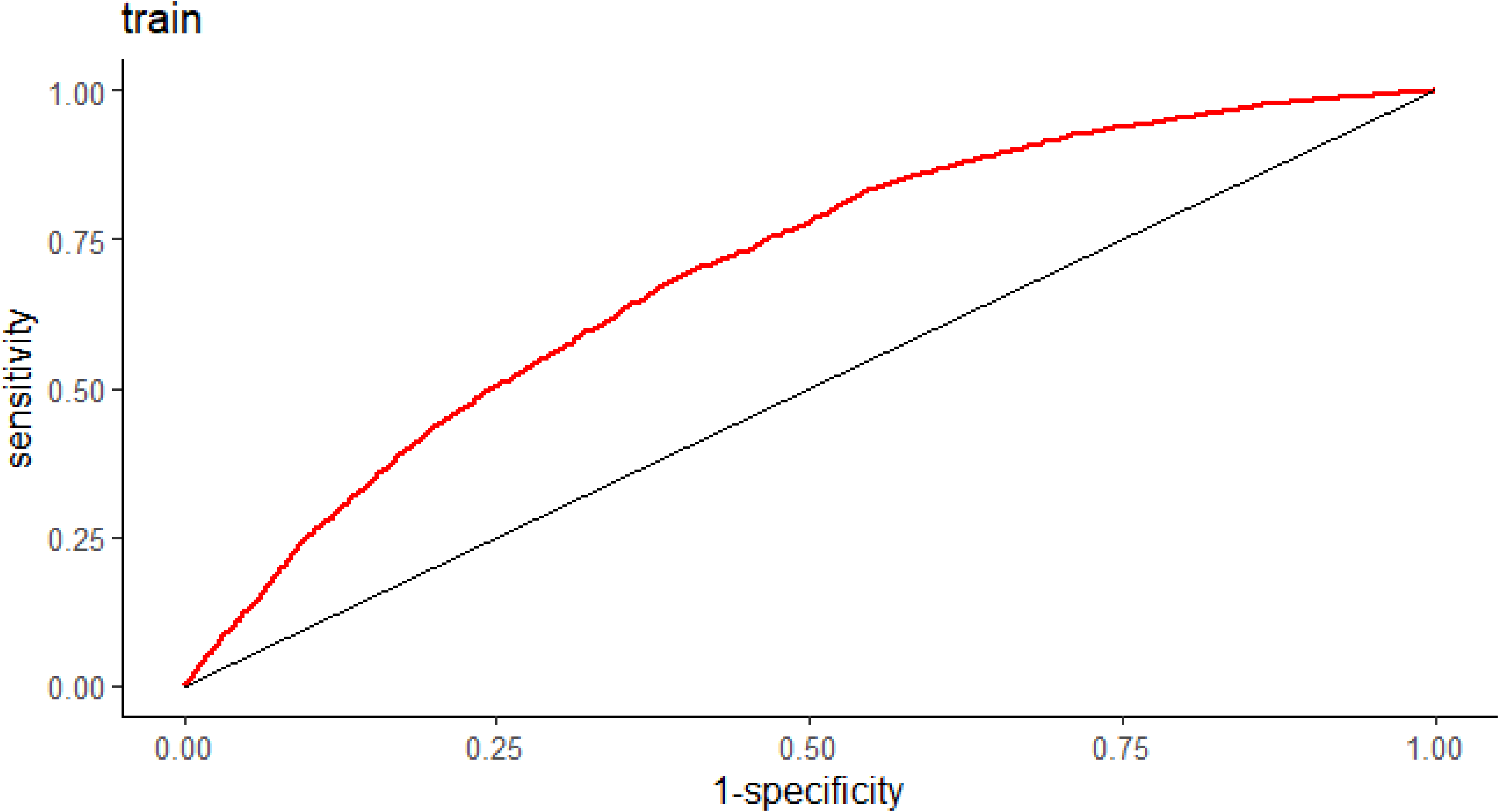
The ROC curve of the training set in nomogram 2.

**Figure 11.**
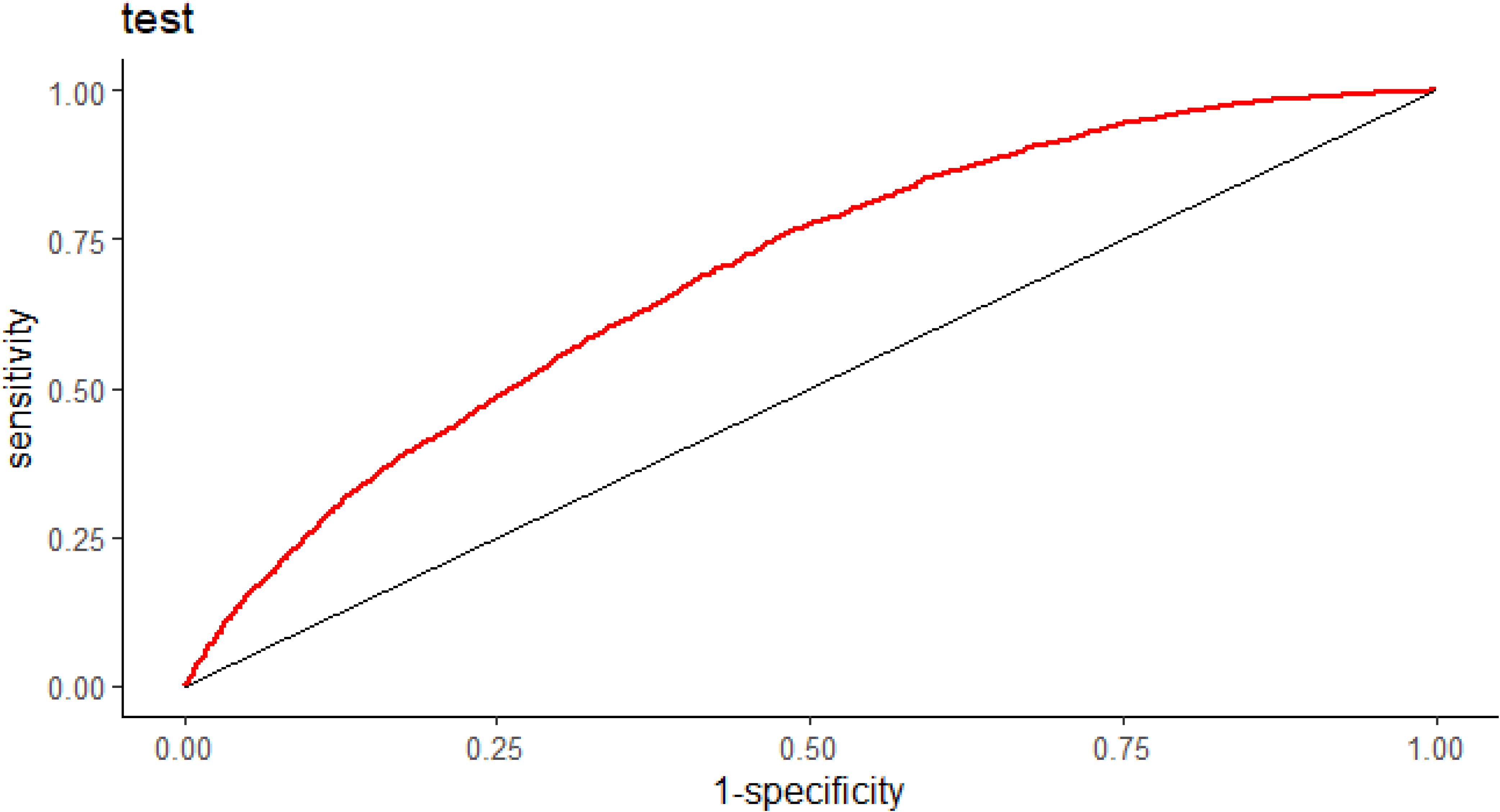
The ROC curve of the validation set in nomogram 2.

Figure 12 and 13 show that the higher the threshold values are, the greater the net benefit of the nomograms decision curves. This indicates the good clinical values of our nomograms.

**Figure 12.**
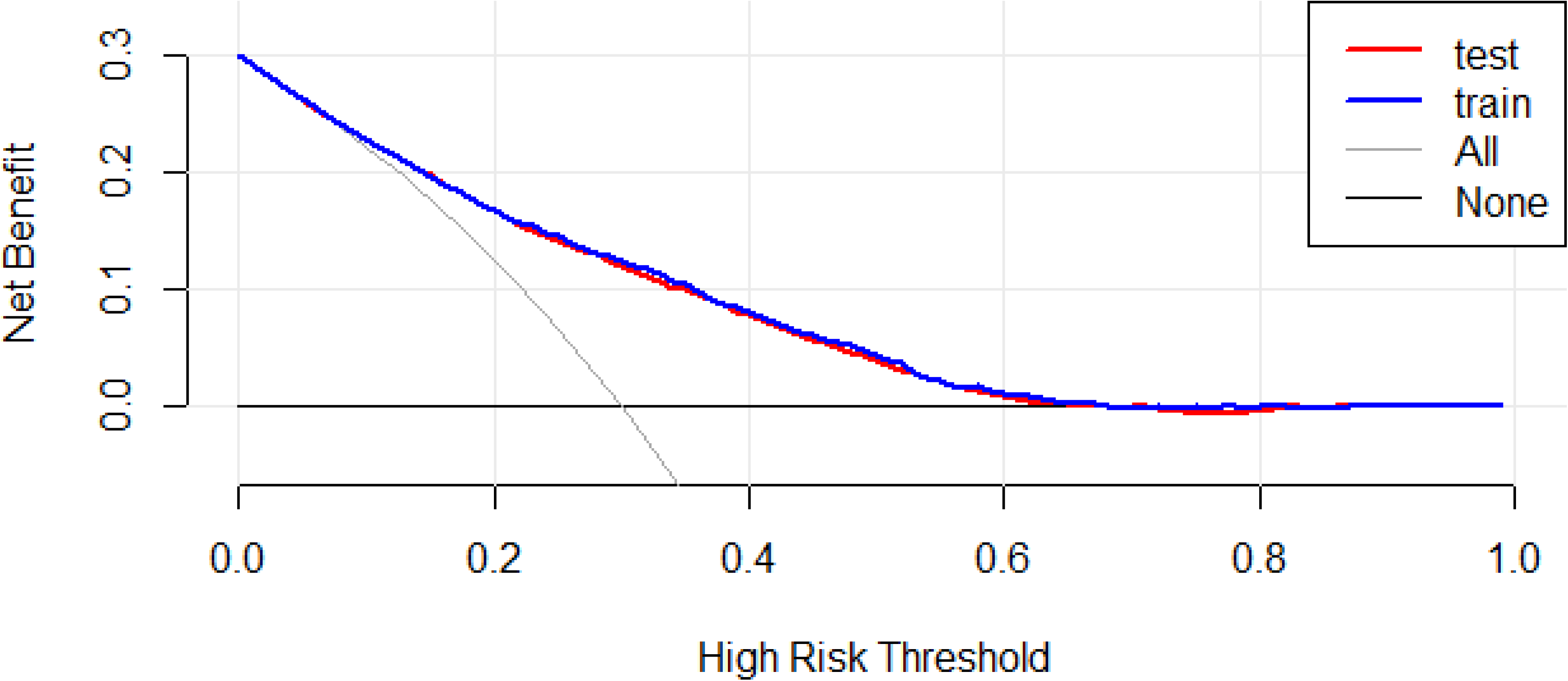
The decision curve analysis of nomogram 1.

**Figure 13.**
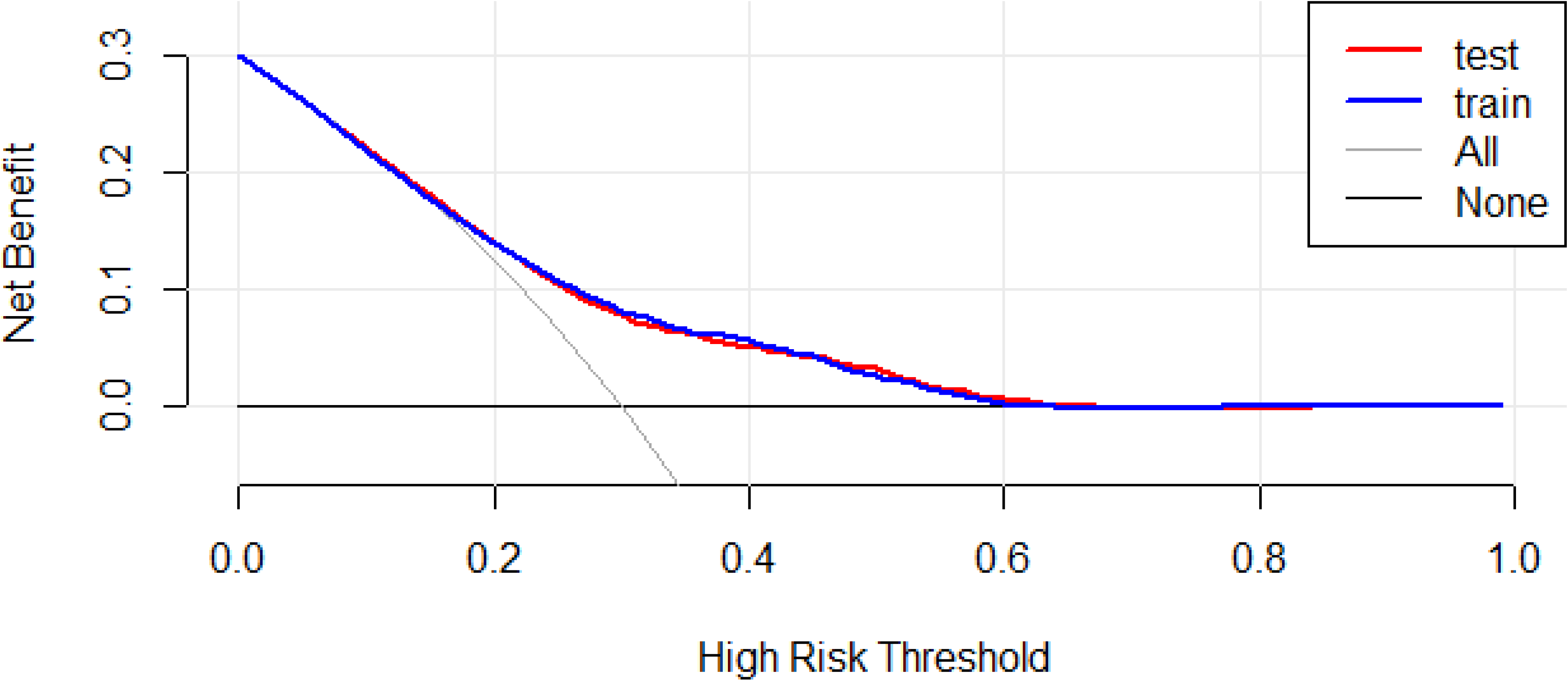
The decision curve analysis of nomogram 2.

## Discussion

This population-based study retrospectively investigated 18,179 ICU patients with AF. Two nomograms were developed and validated to gauge the risk of AHF in those patients. The nomograms contained 10 variables: age; respiratory rate; fluid management; mean corpuscular haemoglobin concentration (MCHC); bicarbonate; blood urea nitrogen (BUN); chloride; and the Charlson, LODS, and SAPS II scores. Both nomograms had good clinical performance. ROC analysis of the training and validation cohorts revealed AUCs of 0.768 and 0.763, respectively, for nomogram 1; these AUCs were 0.696 and 0.692 for nomogram 2. The findings suggest that our models are sufficiently predictive.

The relationship between AF and heart failure remains an area of study [28]. Ischemia, blood pressure, blood glucose, age, and atherosclerosis are generally regarded as risk factors for both diseases [29–31, 33]. The incidences of heart failure and AF both gradually increase with age [32, 33], consistent with our findings.

When heart failure occurs, the respiratory rate increases. Breathing adjustments affect the treatment of heart failure [34–36]. This study recognised respiratory rate as a risk factor for AHF in patients with AF, similar to the results in some previous studies [37]. Input and output volumes are important for evaluating the degree of heart failure [38, 39]; reductions in output and input are termed fluid management. Our model 1 showed that greater fluid management increases the risk of AHF. Relative erythrocyte hypochromia, which manifests as a low MCHC, affects the onset of AHF [40, 41]; a decrease in the MCHC corresponds to an increase in the risk of AHF. The serum bicarbonate level also significantly increases in patients with AHF [42], and it can serve as a predictive indicator. BUN is related to renal function and is not an independent risk factor for AHF, although it can serve as a warning signal [43]. Chloride is associated with cardiorenal and neuroendocrine systems [44–46], and many studies regard the chloride level as a prognostic marker [44]. The present study identified it as a predictive factor, such that a decrease in chlorine was accompanied by an increase in the incidence of AHF. Additionally, the present study utilised three scoring systems to establish the second nomogram. The use of model 2 requires some medical expertise; thus, it is designed for clinicians. Although nomogram 2 is less accurate and predictive compared with nomogram 1, it can provide validation for clinical decision-making.

This study had some limitations. First, all data was obtained from the MIMIC-IV database. Incomplete records and possible data errors may have reduced model accuracy. Second, this was a single-centre retrospective study with internal validation alone; external validation is needed to confirm the reliability of the results. Third, blood lipids were not included in the study due to the lack of corresponding records in the MIMIC-IV database. Blood lipids are important risk factors for cardiovascular diseases [47], and further research is needed concerning their implications.

Fourth, the types and severity of AF were not considered; thus, we did not determine the risk of AHF at each AF severity level. Additionally, all types of AHF were pooled in our analyses. Finally, our nomograms may predict the possibility of AHF in patients with AF, but they cannot determine whether interventions should be promptly applied.

## Conclusion

Two nomograms to predict the risk of AHF in AF patients were developed using data from the MIMIC-IV database. The models had good predictive ability. The nomograms can help to screen AF patients for the risk of AHF and could facilitate good clinical decisions.

## Data Availability

All data generated or analysed during this study are included in this published article and its supplementary information files.

## List of abbreviations

AHF: acute heart failure
AF: atrial fibrillation
MIMIC: Medical Information Mart for Intensive Care
LASSO: least absolute shrinkage and selection operator
AUC: area under the curve
ROC: receiver operating characteristic
DCA: decision curve analysis
C-index: Concordance index
SD: standard deviation
IQR: interquartile range
RBC: red blood cells
WBC: white blood cells
MCV: mean corpuscular volume
RDW: red blood cell distribution width
BUN: blood urea nitrogen
INR: international normalized ratio
PT: prothrombin time
PTT: partial thromboplastin time
RDW: red blood cell distribution width
MCH: mean corpsular hemoglobin
MCHC: mean corpuscular hemoglobin concentration
BMI: body mass index
RAAS: renin angiotensin aldosterone system

## Acknowledgements

We would like to thank all MIMIC project team members.

## Declarations

## Ethics approval and consent to participate

This study is a retrospective analysis of data stored in a public database. All information is available and free for the public, so the agreement of the medical ethics committee board and the need for informed consent were not necessary.

## Consent for publication

Not applicable.

## Supplementary materials

**Figure 14.**
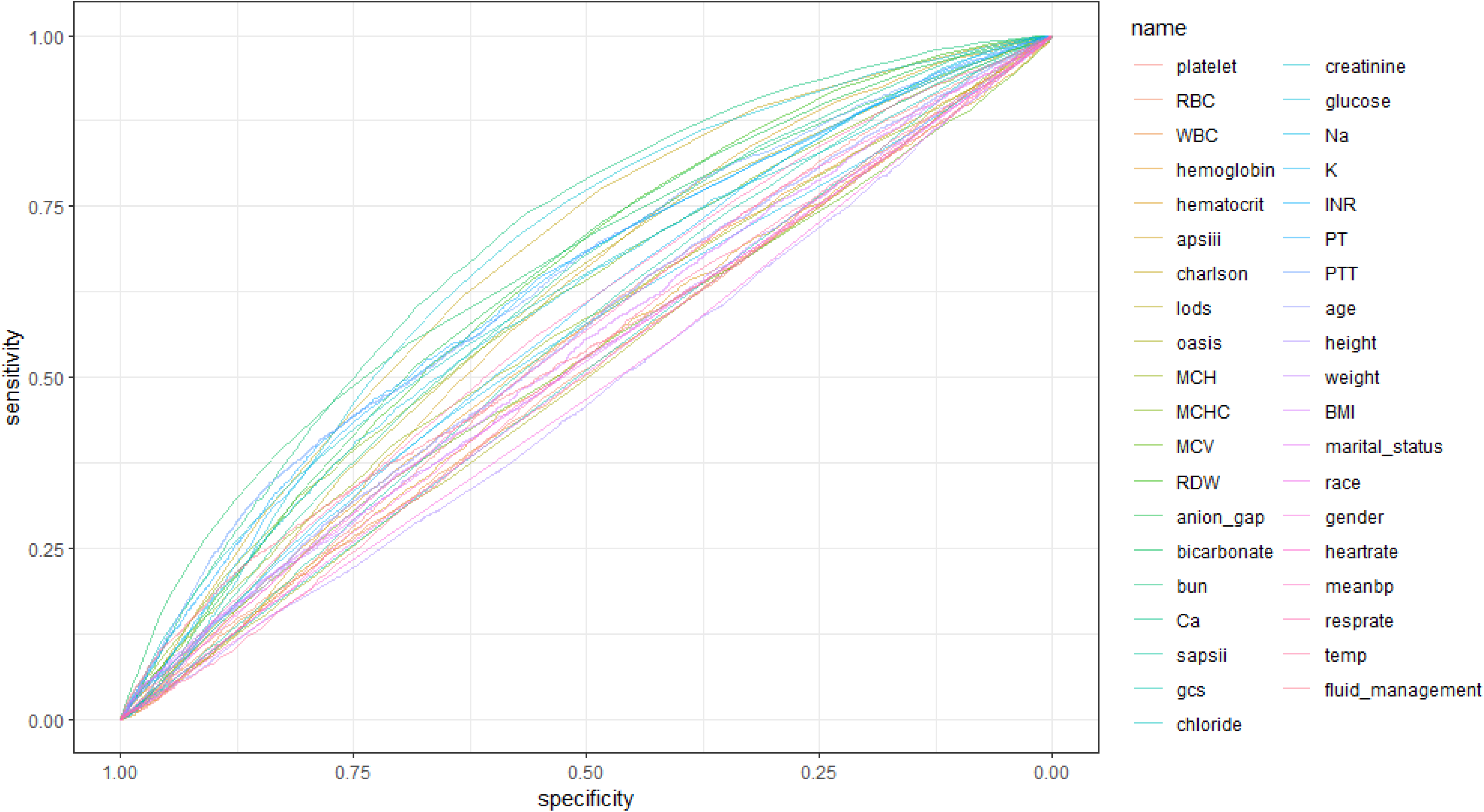
ROC curves of all screened variables.

**Table 2.** AUC values of all screened variables.

|  |  |
| --- | --- |
| bun.auc | 0.693723 |
| creatinine.: | 0.667555 |
| charlson.au | 0.666832 |
| bicarbonat | 0.6574 |
| RDW.auc | 0.637169 |
| anion_gap. | 0.636693 |
| PTT.auc | 0.635668 |
| PT.auc | 0.630236 |
| INR.auc | 0.623605 |
| Ca.auc | 0.613499 |
| chloride.au | 0.612968 |
| glucose.auc | 0.606104 |
| MCHC.auc | 0.60373 |
| lods.auc | 0.602316 |
| apsiii.auc | 0.596075 |
| K.auc | 0.577893 |
| resprate.au | 0.575868 |
| MCH.auc | 0.56197 |
| sapsii.auc | 0.55361 |
| age.auc | 0.553435 |
| Na.auc | 0.548635 |
| hemoglobii | 0.548481 |
| meanbp.au | 0.547079 |
| platelet.au | 0.544108 |
| BMI.auc | 0.541347 |
| fluid_man | 0.539073 |
| weight.auc | 0.524615 |
| MCV.auc | 0.519201 |
| marital_sta | 0.518705 |
| hematocrit | 0.514507 |
| heartrate.a | 0.513549 |
| gcs.auc | 0.510718 |
| temp.auc | 0.510425 |
| WBC.auc | 0.509865 |
| RBC.auc | 0.509729 |
| race.auc | 0.504742 |
| oasis.auc | 0.501189 |
| gender.auc | 0.481012 |
| height.auc | 0.474853 |

**Figure 15.**
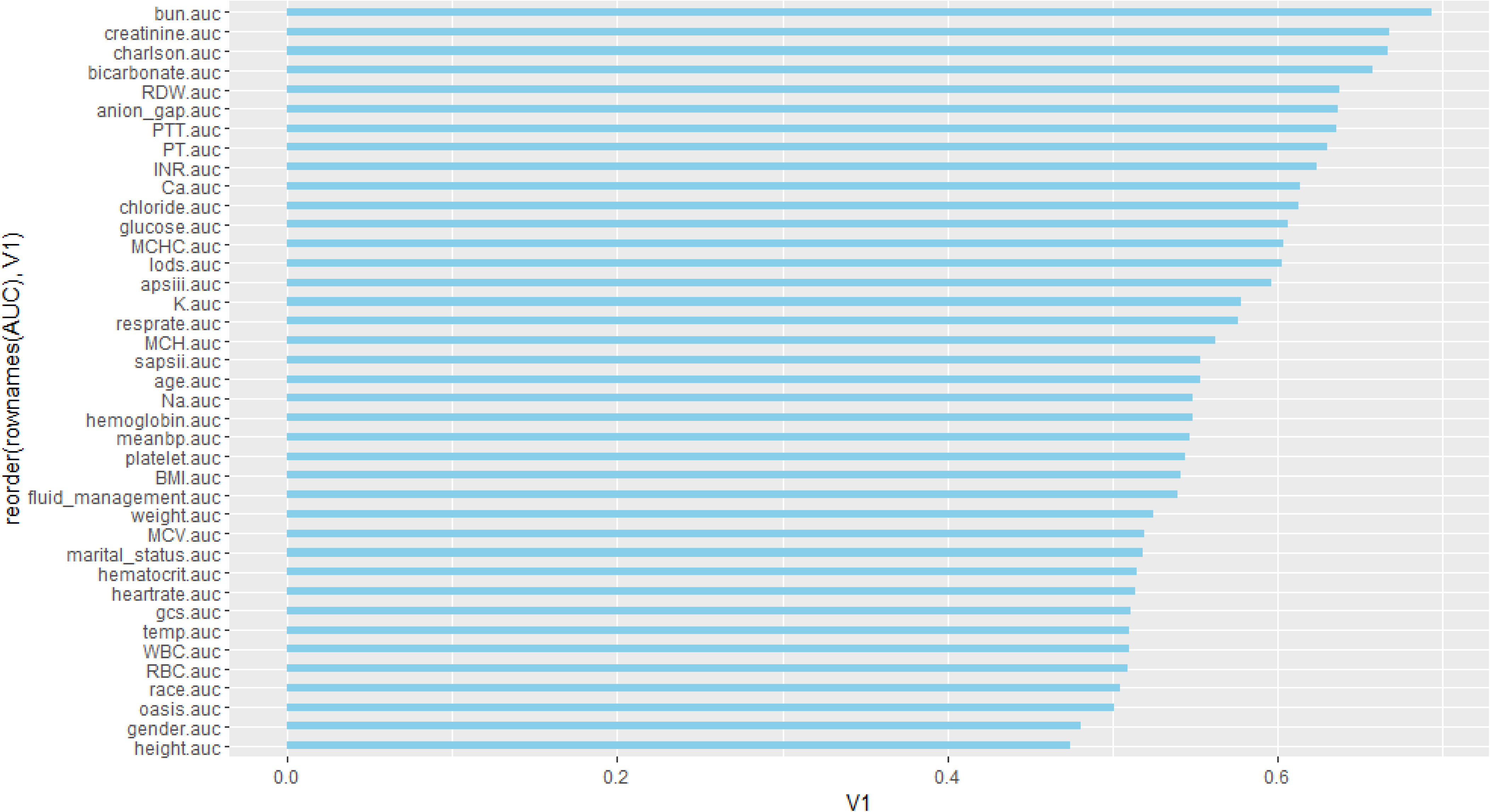
AUC histogram involving all screened variables.

**Figure 16.**
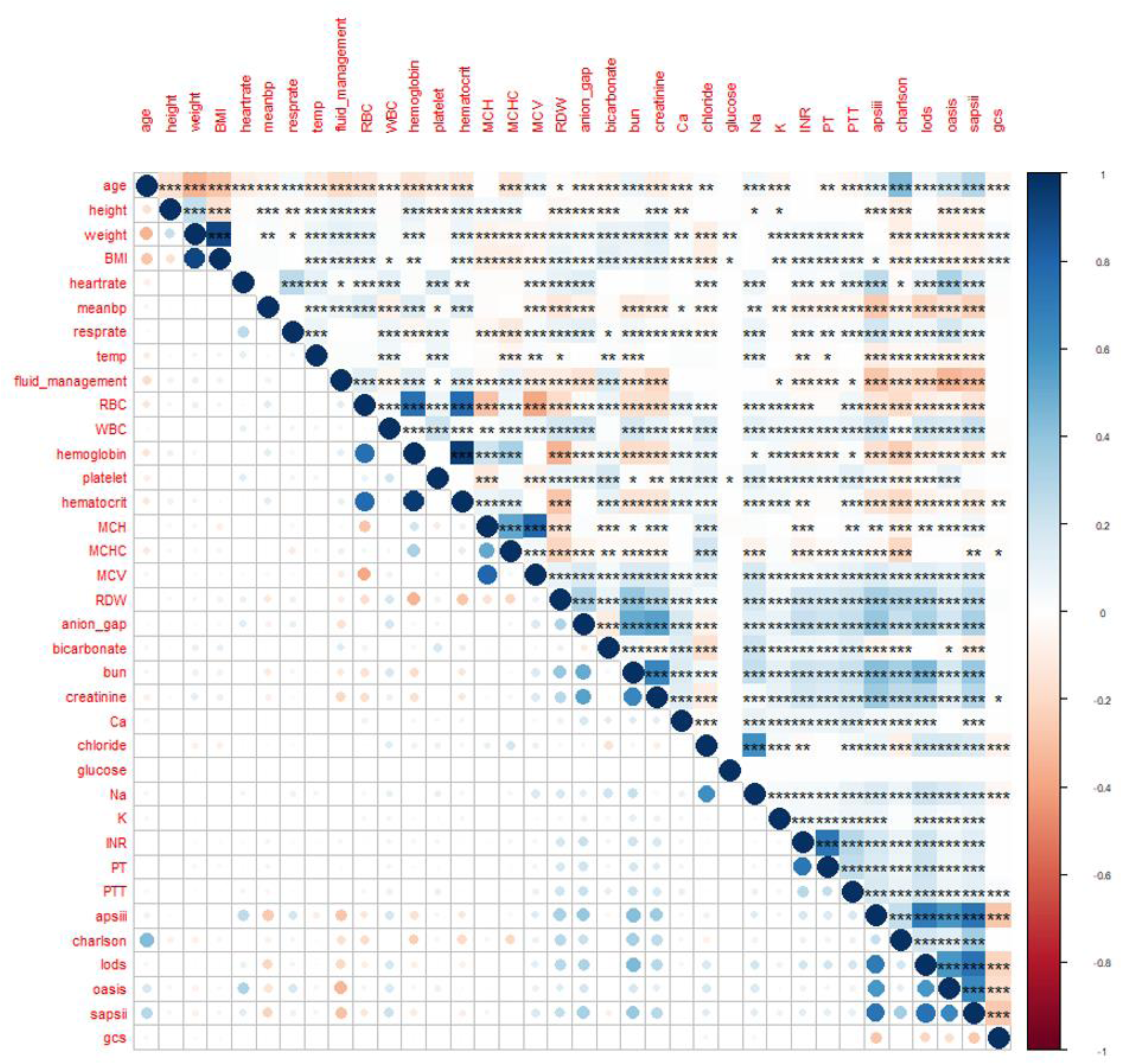
Heatmap involving all screened variables.

## Author contributions

Ziyang Wu conceived, designed the study and drafted the manuscript. Yong Qiao, Gaoliang Yan, Yuhan Qin, Huihong Tang and Shiqi Liu drafted the manuscript. Chengchun Tang and Dong Wang revised the manuscript and approved the final version to be published. All authors read and approved the final manuscript.

The English in this document has been checked by at least two professional editors, both native speakers of English. For a certificate, please see: http://www.textcheck.com/certificate/aTgAHx

